# Altered Glutamate Homeostasis in Paramagnetic Rim Lesions of Patients with Multiple Sclerosis

**DOI:** 10.64898/2026.08.06.26359875

**Authors:** Paul S. Jacobs, Bailey Spangler, Nikka Bakhtiar, Ahmed Elkady, Neil E. Wilson, Anshuman Swain, Elizabeth A. Horwath, Marina M. Awad, Luana Yamashita, Russell T. Shinohara, Simon Thebault, Amit Bar-Or, John A. Detre, David A. Rudko, Matthew K. Schindler, Ravinder Reddy

## Abstract

Paramagnetic rim lesions are a subset of focal white matter lesions specific to multiple sclerosis that are chronically inflamed and are associated with increased tissue injury, brain atrophy, and clinical disability. The molecular mechanisms linking paramagnetic rim lesions to these progressive biological outcomes remain unclear. Glutamatergic dysregulation has been hypothesized as a mechanism of multiple sclerosis progression potentially via excitotoxicity, but lesion-specific involvement is unknown. Here, 7T MRI was used to investigate glutamate-related metabolic alterations in paramagnetic rim lesions. Glutamate-weighted chemical exchange saturation transfer, together with T_1_ mapping and quantitative susceptibility mapping, was evaluated across paramagnetic rim lesions, non-paramagnetic rim lesions, and normal-appearing tissues in participants with multiple sclerosis (n=20) and healthy controls (n=11). Glutamate-weighted chemical exchange saturation transfer contrast was significantly higher in paramagnetic rim lesions compared to non-paramagnetic rim lesions (+10.7%) and normal-appearing white matter (+13%), while no differences were observed in normal-appearing tissue between multiple sclerosis and healthy controls. Additionally, reduced glutamate-weighted chemical exchange saturation transfer contrast in normal-appearing tissues was associated with worse motor and dexterity performance, linking observed metabolic abnormalities to clinical disability. These results identify a distinct metabolic phenotype of paramagnetic rim lesions marked by elevated glutamate-weighted signal consistent with localized excitotoxic stress. This work also implicates lesion-specific glutamatergic dysregulation in paramagnetic rim lesion-related neurodegeneration and demonstrates the potential of metabolic MRI to probe pathogenic mechanisms in multiple sclerosis.

## INTRODUCTION

Best known as an excitatory neurotransmitter, glutamate plays many significant roles in normal brain function and is also an important immune modulator^1^. Dysregulation of glutamate has been implicated in many neurological diseases, including multiple sclerosis, an inflammatory and neurodegenerative disease affecting the central nervous system (CNS). Excess glutamate can induce excitotoxicity leading to tissue injury. Under normal conditions, glutamate is released into the synaptic space^2^, where it is taken up by astrocytes through excitatory amino acid transporters (EAATs), converted into glutamine, and then recycled back to neurons, in part, to prevent excitotoxicity^3^. It has been suggested that, in multiple sclerosis, this recycling process may be disrupted by excess glutamate production from immune cells and reduced astrocytic uptake^4^, resulting in excitotoxic neuronal death. In the white matter, oligodendrocytes also mediate glutamate recycling, and the loss of these cells within demyelinating multiple sclerosis lesions could further contribute to both acute and chronic injury, with consequent progressive axonal neurodegeneration^5–7^. Previous studies investigating glutamate regulation within mouse models of multiple sclerosis have shown increased concentrations of hippocampal glutamate compared to wild type mice^8,9^. Furthermore, histopathology studies of multiple sclerosis brain have also shown increased levels of proteins involved in glutamate homeostasis in active lesions compared to normal white matter from healthy controls^10^, supporting the hypothesis that glutamatergic dysregulation plays an important role in multiple sclerosis pathophysiology.

Paramagnetic rim lesions (PRLs) are an emerging MRI marker associated with progressive neurodegeneration and worse clinical disability in multiple sclerosis^11–13^. PRLs are a subset of focal white matter lesions, and their histopathologic correlate is the chronic-active lesion (CAL), whose characteristic rim of chronic inflammation is due to iron-laden microglia^14^. PRLs are best identified on susceptibility-weighted MRI and defined by a hypointense rim seen in T * phase images, or hyperintense rim on quantitative susceptibility mapping (QSM). PRLs are associated with more severe tissue loss compared to non-PRLs and increased prevalence of PRLs is associated with greater brain atrophy and earlier more severe clinical disability accumulation^15^. However, the mechanisms linking PRLs to progressive tissue loss remains unclear, and whether glutamatergic dysregulation mediates this relationship is currently not known.

*In vivo* measurements of CNS glutamate are limited. ^1^H MR spectroscopy (^1^H MRS) allows for the non-invasive measurement of several brain metabolites (including glutamate). MRS research studies in multiple sclerosis have reported increased glutamate + glutamine (Glx) in both acute, contrast-enhancing lesions and normal appearing white matter (NAWM)^3^, while other studies reported a 20% decrease in Glx in the normal appearing gray matter (NAGM)^16^ and decreased Glx in the hippocampus^17^ in participants with multiple sclerosis (PwMS) compared to healthy controls. A major limitation of these previous studies was that they were all performed at 3T. Lower field strength systems do not provide adequate chemical shift dispersion to resolve glutamate and glutamine peaks. Additionally, they have lower spatial resolution, leading to partial volume averaging. More recently, Swanberg et al.^18^ acquired MRS at 7T and reported a decrease in glutamate in the frontal cortex of PwMS compared to healthy controls.

To overcome some of the limitations of MRS, chemical exchange saturation transfer (CEST) techniques can image certain metabolites containing labile protons, using MRI with improved spatial resolution. In CEST MRI, frequency-selective saturation is applied to the exchangeable protons on a given metabolite. This saturated magnetization then exchanges with the surrounding bulk water causing a decrease in the bulk water signal of the tissue. The glutamate’s amine protons exchange at a rate which allows it to be imaged at 7T using a flavor of this technique called glutamate-weighted CEST (GluCEST)^19^. Previous experiments have shown that approximately 80% of the GluCEST signal originates from the glutamate pool while various other macromolecules contribute to the rest of the signal^20^. The improved spatial resolution allows for quantification of GluCEST contrast within smaller structures including multiple sclerosis lesions (i.e., PRLs and non-PRLs). O’Grady et al.^21^ performed GluCEST acquisitions on a cohort of PwRRMS and reported greater GluCEST contrast in the prefrontal cortex NAGM of PwMS compared to healthy participants. However, in a later study using glutamate-weighted apparent exchange-dependent relaxation (AREX), the same group found no contrast changes in NAGM or NAWM compared to healthy controls^22^. While these previous 3T MRS and 7T GluCEST imaging studies found variable changes in Glx and/or glutamate, these studies did not assess for changes in signal within or associated with PRLs.

Therefore, the primary aim of this work is to employ GluCEST imaging at 7T to quantify differences in relative glutamate-weighted MR contrast between PRLs and non-PRLs, normal appearing brain tissue types (NAGM and NAWM), and clinical disability measures in a cohort of PwMS compared to healthy controls. Relationships between GluCEST contrast and clinical measures of disability are also investigated. Additionally, considering that prior literature has primarily focused on QSM and T_1_ relaxation mapping in the context of multiple sclerosis disease progression and more specifically PRL formation, GluCEST imaging results will be compared to these more commonly utilized types of acquisitions.

## SUBJECTS/MATERIALS AND METHODS

### Participants and Clinical Measures

All data were acquired after obtaining written informed consent under an approved University of Pennsylvania Institutional Review Board research protocol. MS participants were recruited between October 2023 and December 2024 from the University of Pennsylvania’s Department of Neurology, Multiple Sclerosis and Related Disorders Outpatient Clinic. 7T MRI images were acquired on 20 participants with MS (13 female, 7 male; 17 RRMS, 2 radiologically isolated syndrome (RIS), 1 primary progressive (PPMS)) and 11 healthy participants (7 female, 4 male). The average age of the MS cohort was 39.1 years (range of 21-64 years), and the average age of the healthy participants was 35.0 years (26-45 years). When evaluated using a t-test, no statistically significant difference was observed in the age distribution between the two groups (*P* = 0.30). Four MS participants were not on disease modifying therapies (DMT) at the time of scanning,15 participants were on anti-CD20 DMTs and 1 participant was on cladribine. Disease duration was defined as the amount of time since the onset of clinical symptoms attributable to MS, or first observation of MRI findings indicative of MS, which was on average 7.4 years. Demographic and clinical disease metrics described here can also be seen summarized in Table 1. All MS participants met the 2024 McDonald’s diagnostic criteria^23^. In 15 of the 20 participants with MS, 2 quantitative clinical measures of function were collected during pre-imaging assessments on MS participants, which included timed 25-foot walk test (T25FW) and 9-hole peg test (9-HPT).

**Table 1.**
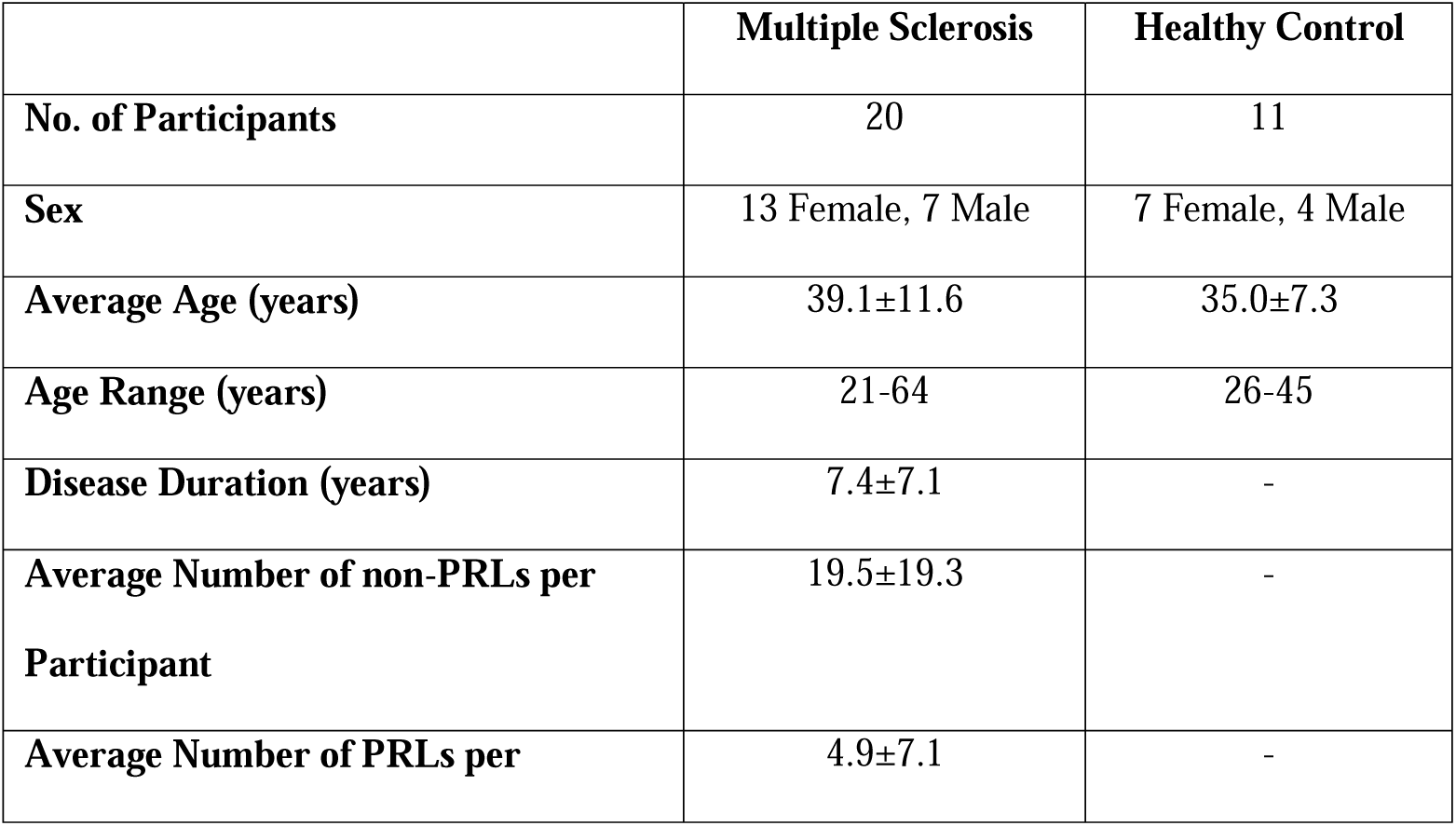
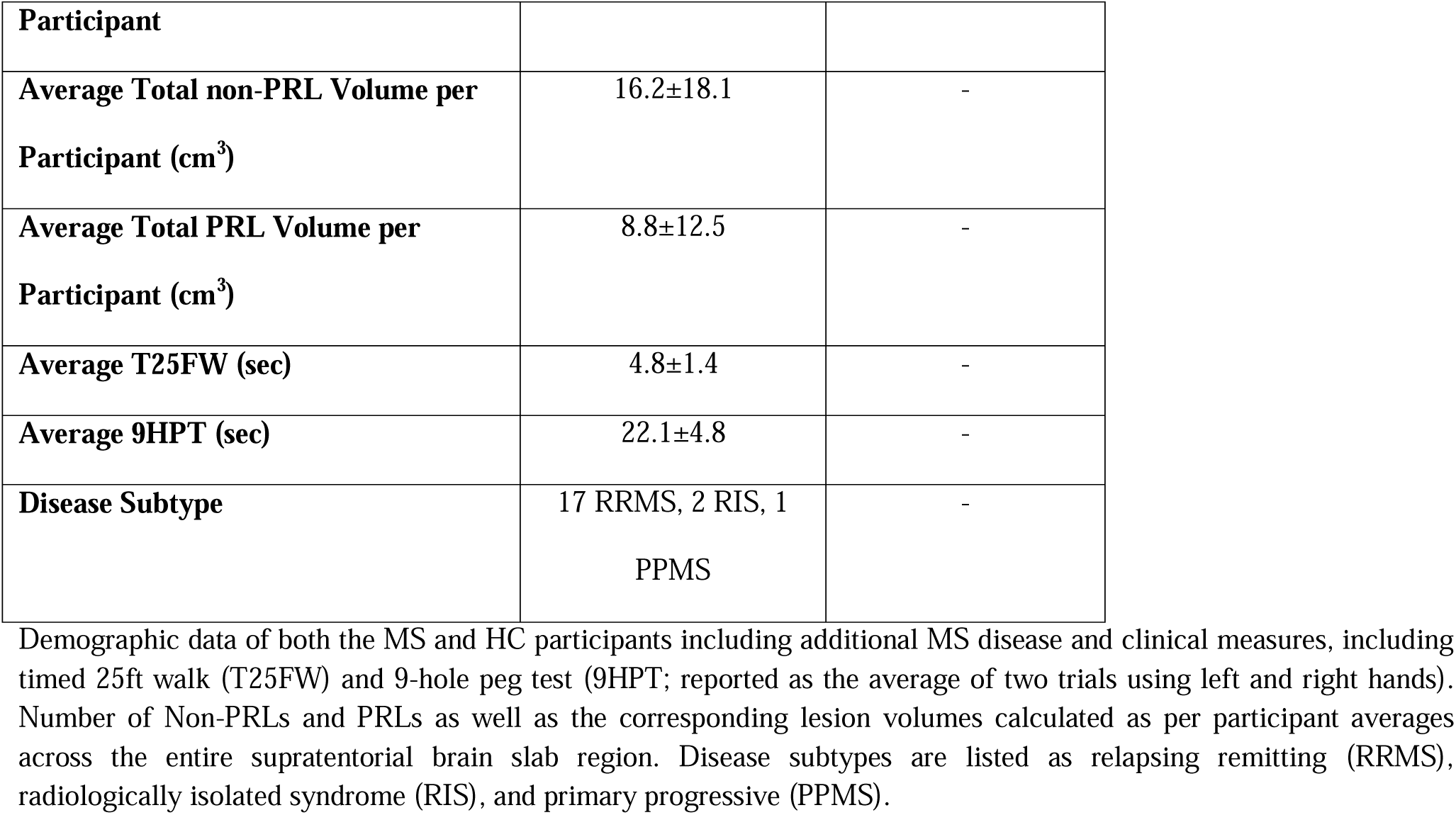
Summarized Patient Demographic Data.

### MR Image Acquisition

All participant images were acquired on a 7T MRI system (MAGNETOM Terra, Siemens Healthcare, Erlangen, Germany) using a single-channel transmit / 32-channel receive phased-array head coil (Nova Medical, Wilmington, MA, USA). GluCEST imaging was performed with a modified turbo FLASH sequence, as previously described in Cai et al.^19^, using a TR = 3.5 ms, TE = 1.79 ms, and shot TR = 6000 ms. GRAPPA was applied with an acceleration factor of 2. Magnetization preparation was achieved using a B_1,rms_ of 3.06 µT, and saturation durations of 800 ms (eight 99 ms Hanning shaped pulses with 1 ms interpulse delay) were applied at offset frequencies of ±1.8 to ±4.2 ppm in step sizes of 0.3 ppm. The 3D acquisition volume had a thickness of 24 mm (12 slices, 2 mm slice thickness) with an in-plane resolution of 1 × 1 mm^2^ and a matrix size of 240 × 196. In this acquisition each slice was acquired in three shots. In the case of MS participants, the field of view was generally placed in a slight oblique orientation in a region with a high number of lesions, typically near the corpus callosum, while in HC participants the volume was placed in a consistent axial orientation just superior to the corpus callosum.

Using the same spatial parameters as the GluCEST slab the following sequences were also acquired: 1) Water saturation shifting referencing (WASSR) images^24^: -1.5 to 1.5 ppm (step-size of 0.15 ppm) with a pulse saturation magnitude of B_1,rms_ = 0.29 µT and a 200 ms duration. WASSR images were utilized to generate B_0_ maps. 2) MP2-RAGE^25^ to generate T_1_ maps: TR = 4.60 ms, TE = 2.19 ms, shot TR = 5000 ms, TI_1_ = 700 ms, TI_2_ = 2700 ms. 3) B_1_^+^ maps were generated as described in Volz et al.^26^, using magnetization preparation pulses having flip-angles = 20°, 40°, and 80°. A second set of whole brain MP2-RAGE images were acquired with the same parameters as previously described with the following changes: isotropic resolution of 0.70 mm^3^, matrix size = 396 × 352, and number of slices = 224. Whole brain FLAIR images were also acquired using a modified SPACE sequence^27^ with the following parameters: isotropic resolution of 0.75 mm^3^, TR = 7600 ms, TI = 2400 ms, TE = 476 ms, flip-angle = 120°, turbo factor = 230, matrix size = 320 × 320, and number of slices = 208. For the purposes of confirmatory PRL identification, a 3D high-resolution T_2_*-weighted multi-echo gradient recalled echo (ME-GRE) image set was acquired over a 24 mm volume (directly matching the volume and orientation of the GluCEST acquisition) with the following parameters: in-plane resolution of 0.21 × 0.21 mm^2^, slice thickness = 1 mm, TR = 1320 ms, TE_1_ = 15 ms, TE_2_ = 32 ms (2-echos), flip-angle = 50°, matrix size = 768 × 1024, and number of slices = 24. To generate the QSM images, 3D whole brain ME-GRE images were obtained using the following parameters: isotropic resolution of 0.80 mm^3^, TR = 40 ms, TE_1_ = 6 ms, TE_2_ = 12 ms, TE_3_ = 18 ms, TE_4_ = 24 ms, TE_5_ = 30 ms, TE_6_ = 36 ms (6-echos), flip-angle = 13°, matrix size = 252 × 324, and number of slices = 128.

### Image Post-processing and Analysis

CEST-weighted images were interpolated to a second order polynomial and evaluated at +/- 3ppm accounting for B_0_ off resonance effects. B_1_ correction was performed in two steps, first calcium titanate dielectric pads (7TNS, Multiwave Imaging, Marseille, France) were placed on either side of the participant’s head (near the temporal lobes) during acquisition which aided in increasing the overall B_1_ field homogeneity and magnitude^28^ and second, a post-processing correction was applied as described in Cember et al.^29^. GluCEST contrast was calculated via asymmetry analysis between the 3ppm and -3ppm frequency offsets, as seen in Eq. 1 below. It should be noted that prior to quantification the first and last imaging slices were discarded due to phase artifacts.

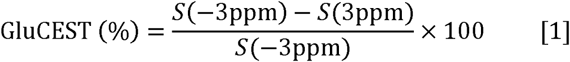

Lesion masks were manually generated (BS, NB, MKS) on the 3D whole brain MP2-RAGE for every MS participant using ITK-SNAP^30^. Initial PRL identification was performed on unwrapped phase images from the high-resolution T *-weighted ME-GRE acquisition. Lesions were then visually identified on the unwrapped 0.80 mm^3^ isotropic resolution ME-GRE phase images averaged from the 3^rd^, 4^th^, and 5^th^ echo times by a clinician with 10 years of experience in identifying PRLs (MKS) which were classified based on the NAIMS criteria^13^. Non-PRL tissue was masked separately from PRL tissue, and all image contrast quantifications were made over the registered 20 mm image acquisition slab of interest, which we will refer to in the rest of the work as the “brain slab” region. By referencing both these lesion segmentations and the anatomical images from the acquisition region, the number of PRLs and non-PRLs within the imaging slab were quantified. As a note, lesions that were highly diffuse in appearance were counted as a single lesion. A representative example of the size and location of this acquired brain slab region can be seen in Supplement Figure 1. While the size of the 3D acquisition region remained constant across the participants the location and orientation did vary in order to capture lesions of interest. Lesion boundaries were masked using the T_1_ image from the MP2-RAGE acquisition. T_1_ maps were used to generate gray and white matter segmentation masks for a separate tissue-based quantification and NAGM and NAWM^31^.

ME-GRE data were separately processed to calculate QSM images by first utilizing the FSL Brain Extraction Tool (BET) to mask the brain with a BET intensity threshold of 0.4 and a smoothing parameter of 0.2^32^. Phase images from the ME-GRE acquisition were unwrapped using a combination of spatial and temporal unwrapping techniques. Spatial unwrapping was performed using Phase Region Expanding Labeler for Unwrapping Discrete Estimates (PRELUDE)^33^. Due to its superior reduction of non-local field effects caused by high-susceptibility brain interfaces and its ability to preserve brain edges, the Quantitative Susceptibility Mapping Artifact Reduction Technique (QSMART)^34^ was used for QSM image generation. QSMART background field removal parameters were applied as follows: fit threshold = (0.5, 0.5, 500), spatial dependent filtering used a vasculature spherical radius of 8 pixels and had default tuning parameters of (σ_c_ = 10, σ_y_ = 2, σ_I_ = 10, and C_s_ = 500)^34,35^. Finally, the iterative least square (iLSQR) method was applied for dipole field inversion^34,36^. It should be noted that QSM images were only generated for the MS participants due to ME-GRE data not being collected for the HC scans.

### Statistical Analysis

Mean and standard deviation values were calculated for GluCEST contrast, T_1_ values, and QSM values within the cerebral tissue brain slab volume (WB), normal appearing gray matter (NAGM), normal appearing white matter (NAWM), and lesion tissue (PRLs and non-PRLs). Two-sample t-tests with a 95% confidence interval were used to compare whole brain, and NAGM contrast values between HC and MS participants. For the GluCEST contrast and T_1_ value comparisons involving multiple tissue types within the MS group (NAWM, Non-PRL, and PRL) and HC (WM), a one-way analysis of variance (ANOVA) was performed for each contrast. Significant ANOVA results were followed by post-hoc pairwise comparisons using the Tukey-Kramer adjustment to account for multiple comparisons. The same groupwise comparisons and multiple comparison adjustment were made for QSM values but only within MS tissue subtypes due to lack of HC participant QSM data. It should also be noted that group wise analyses were exploratory and meant for hypothesis generation. Pearson correlations were performed between the tissue contrasts (GluCEST, T_1_ values, and QSM values) and the following covariates: age, lesion volume, disease duration, and clinical metrics (T25FW & 9-HPT). An additional multiple linear regression was performed to evaluate the effect of lesion type (PRL vs. non-PRL) and lesion volume on GluCEST contrast, including a type-by-volume interaction term. All analyses were conducted using MATLAB (The MathWorks, Natick, MA, USA) and R Statistical Software (v4.3.1), and a significance level of α = 0.05 was used for all statistical tests.

## RESULTS

### Qualitative Image Comparisons Between MS and HC

Representative images including FLAIR, QSM, and GluCEST contrast maps of a HC and two PwMS (MS1 and MS2) are shown in Figure 1. In the PwMS FLAIR images, a large lesion in the white matter (periventricular lesion in MS1 and juxtacortical lesion in MS2) is seen for each subject. MS1’s lesion has a hyperintense rim on QSM that is characteristic of a PRL while MS2’s lesion lacks the characteristic finding of a PRL (i.e. non-PRL). On GluCEST imaging, the PRL is hyperintense throughout the entire lesion (reflecting higher glutamate-related signal) while the non-PRL appears as hypointense (reflecting lower glutamate-related signal) relative to NAWM.

**Figure 1.**
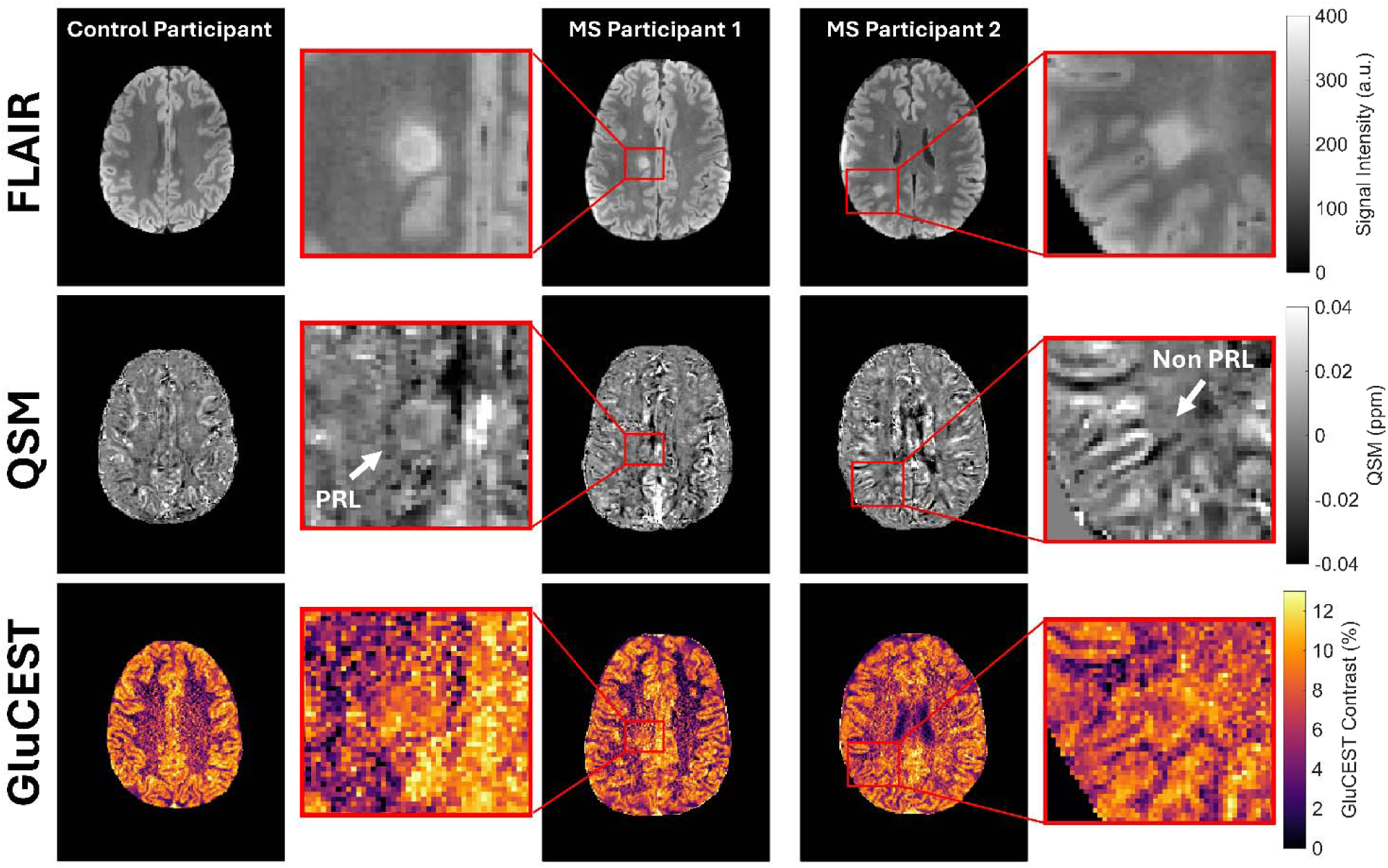
PRLs and Non-PRLs Exhibit Differing Contrasts Compared to Healthy Tissue. A figure showing a qualitative FLAIR, QSM, and GluCEST comparison between a healthy control participant and two MS participants. Zoomed in lesion regions can be seen for each MS participant for all three contrasts in which both FLAIR images appear as hyperintense while the QSM image confirms a distinguishable rim (indicating a PRL) in MS participant 1 and no rim in MS participant 2 (indicating a non-PRL). Similarly, the GluCEST images show the PRL in MS participant 1 as hyperintense, while the non-PRL in MS participant 2 appears as hypointense.

Figure 2 presents a comparison of quantitative and semi-quantitative sequences in four additional PwMS showing MP2-RAGE T_1_ maps, QSM, and GluCEST contrast maps. Zoomed in views of the regions of interest are also present showing more detailed examples of lesion contrast. All four PwMS show large periventricular white matter lesions with increased T_1_ relaxation values relative to the surrounding normal appearing tissue. PwMS 3 and 4 each have a lesion with a hyperintense rim on QSM images which is characteristic of a PRL. The lesions seen in PwMS 5 and 6 do not have a hyperintense rim on QSM image, characteristic of non-PRLs. Increased GluCEST contrast is visualized within the PRLs while decreased GluCEST contrast is visualized in the non-PRLs, compared to the surrounding normal-appearing tissue. It should be noted that in the PRLs, PwMS 3 showed increased GluCEST contrast throughout the entire PRL, while PwMS 4 showed an increasing GluCEST contrast gradient moving towards the edge of the PRL.

**Figure 2.**
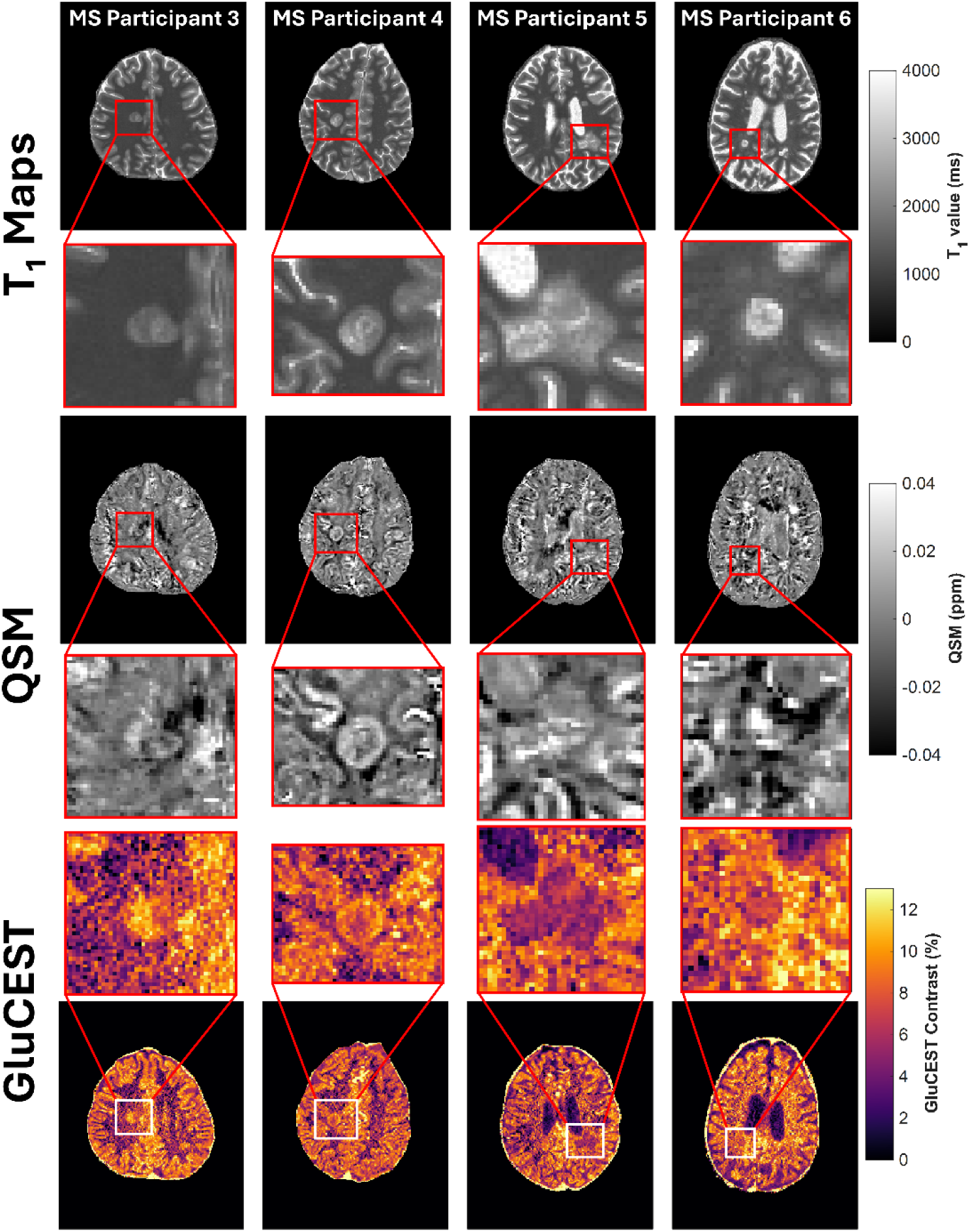
GluCEST Imaging Reveals Increased PRL Contrast Compared to Decreased Non-PRL Contrast. Representative images showing multiple contrasts of lesions in four additional MS participants. (**Rows 1 & 2**) T_1_ maps showing large prominent subcortical (participants 1 & 2) and periventricular (participants 3 & 4) lesions with l nger relaxation times compared to the surrounding NAWM. (**Rows 3 & 4**) QSM images showing the paramagneti rim (participants 1 & 2) and non-paramagnetic rim (participants 3 & 4). (**Rows 5 & 6**) GluCEST maps show that lesions with a visible phase rim also appear to have a higher GluCEST contrast compared to surrounding tissue, while those without a phase rim appear to have lower GluCEST contrast than surrounding tissue.

### Quantitative Image Comparisons Between MS and HC

Results from group wise analysis of GluCEST contrast and T_1_ relaxation times for the brain slab volume and tissue classes (NAGM, NAWM, PRLs, and non-PRLs) comparing PwMS to HCs are shown in Figure 3. GluCEST contrast is greater in PRLs (8.05±0.97%) compared to non-PRLs (7.27±1.36%), a 10.7% increase (*P* = 0.01). GluCEST contrast is also greater in PRLs compared to NAWM (7.12±1.09%), a 13.0% increase (*P* = 0.01). T_1_ relaxation times are longer in PRLs (1653.0±280.9 ms) compared to NAWM (1173.4±22.2 ms), a 40.8% increase (*P* < 0.01) and compared to HC white matter (1155.9±17.1 ms), a 43.0% increase (*P* < 0.01). T_1_ relaxation times are also longer in non-PRLs (1609.2±365.6 ms) compared to NAWM (37.1%) and HC white matter (39.2%). QSM values are increased within PRLs (4.26±7.27 ppb) compared to non-PRLs (-0.68±3.25 ppb), a 720% higher QSM value. QSM values are also 250% higher within PRLs compared to NAWM (-2.77±0.94 ppb) and 180% higher compared to NAGM (2.46±0.95ppb). Additionally, QSM values are also 460% higher compared to non-PRLs. Average GluCEST contrast, T_1_ relaxation times, and QSM values are summarized in Supplemental Table 1.

**Figure 3.**
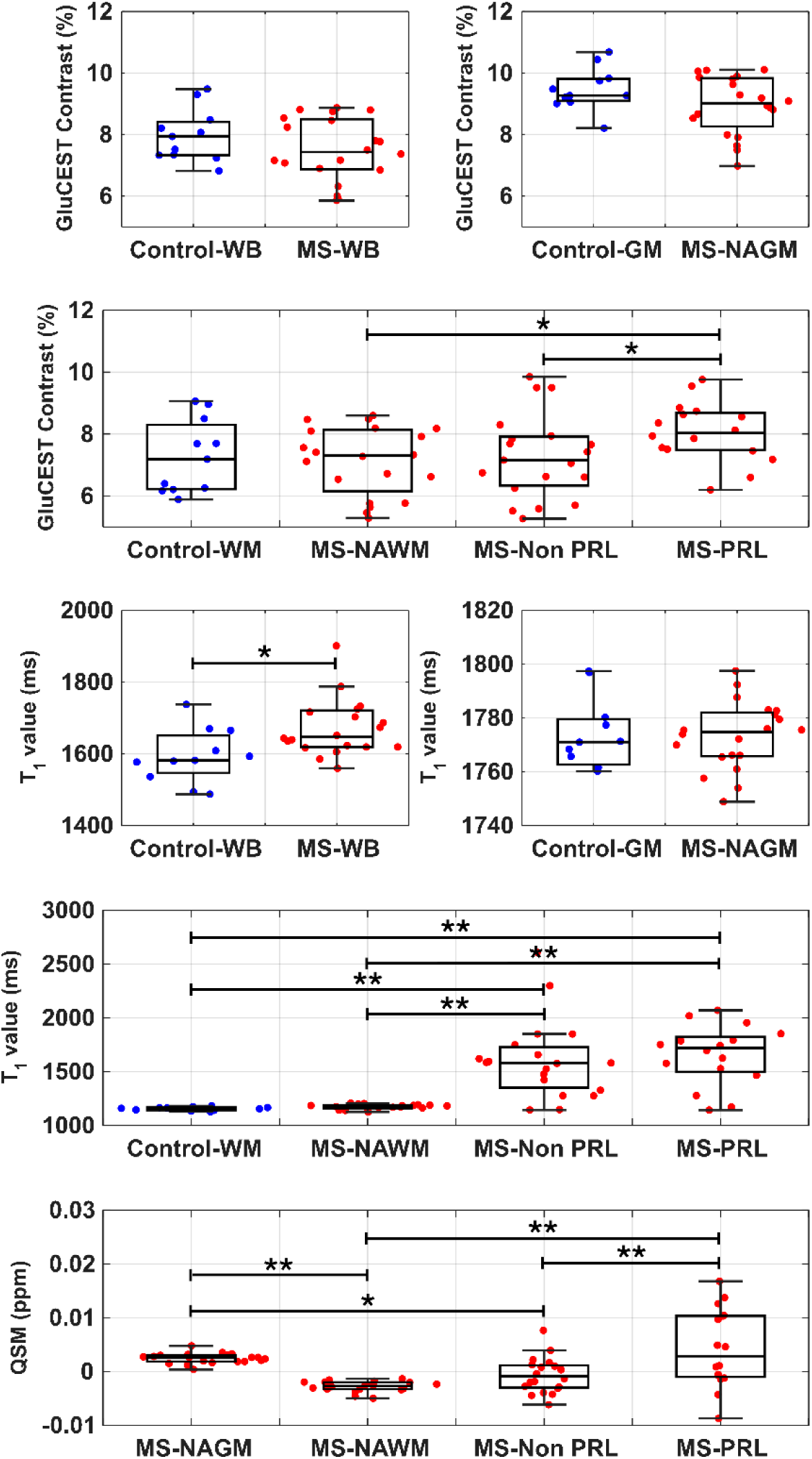
Quantitative Comparison of Image Contrasts Between Tissue Subtypes. Groupwise comparisons of GluCEST contrast values and T_1_ relaxation times in different tissue types between healthy control (blue data points) and MS participants (red data points). For GluCEST values, statistically significant differences (*P* < 0.05 indicated by *, *P* < 0.01 indicated by **) were observed for MS-PRL/MS-NAWM (CI [-2.26, -0.20], *P* < 0.05) and MS-PRL/MS-non-PRL ( CI [-2.12, -0.03], *P* < 0.05). For T_1_ relaxation times, statistically significant differences were observed in the slab volume (t-value = 2.28, CI [9.99, 191.51], *P* = 0.03), MS-PRL/Control-WM (CI [-747.31, -248.75], *P* < 0.01), MS-non-PRL/Control-WM (CI [-694.52, -212.26], *P* < 0.01), MS-PRL/MS-NAWM ( CI [-693.99, -267.05], *P* < 0.01), and MS-non-PRL/MS-NAWM (CI [-639.77, -231.98], *P* < 0.01). QSM value comparisons for different MS tissue types are also shown with statistically significant differences being observed between MS-PRL/MS-non-PRL (CI [-0.008, -0.001], *P* < 0.01), MS-PRL/MS-NAWM (CI [-0.010, -0.003], *P* < 0.01), MS-NAGM/MS-NAWM (, CI [0.002, 0.008], *P* < 0.01), and MS-NAGM/MS-non-PRL (CI [0.001, 0.006], *P* < 0.05).

### Age, Disease Duration, and Lesion Volume Correlations across Imaging Metrics

Analysis of correlations between imaging measures (GluCEST contrast, T_1_ relaxation times, and QSM values) and age is shown in Figure 4. Brain slab and NAGM GluCEST contrast values are both negatively correlated with age (R^2^ = 0.18, *P* = 0.01, R^2^ = 0.18, *P* < 0.01). Brain slab T_1_ relaxation time is positively correlated with age (R^2^ = 0.25, *P* < 0.01) while NAGM T_1_ relaxation time is negatively correlated with age (R^2^ = 0.14, *P* = 0.03). GluCEST and T_1_ are not correlated with age in HCs. Brain slab GluCEST contrast is negatively correlated with disease duration (R^2^ = 0.20, *P* = 0.04). T_1_ relaxation time positively correlates with disease duration across the brain slab volume (R^2^ = 0.32, *P* < 0.01). No significant correlations to disease duration are observed for QSM values. These plots can be seen in Supplemental Figure 2.

**Figure 4.**
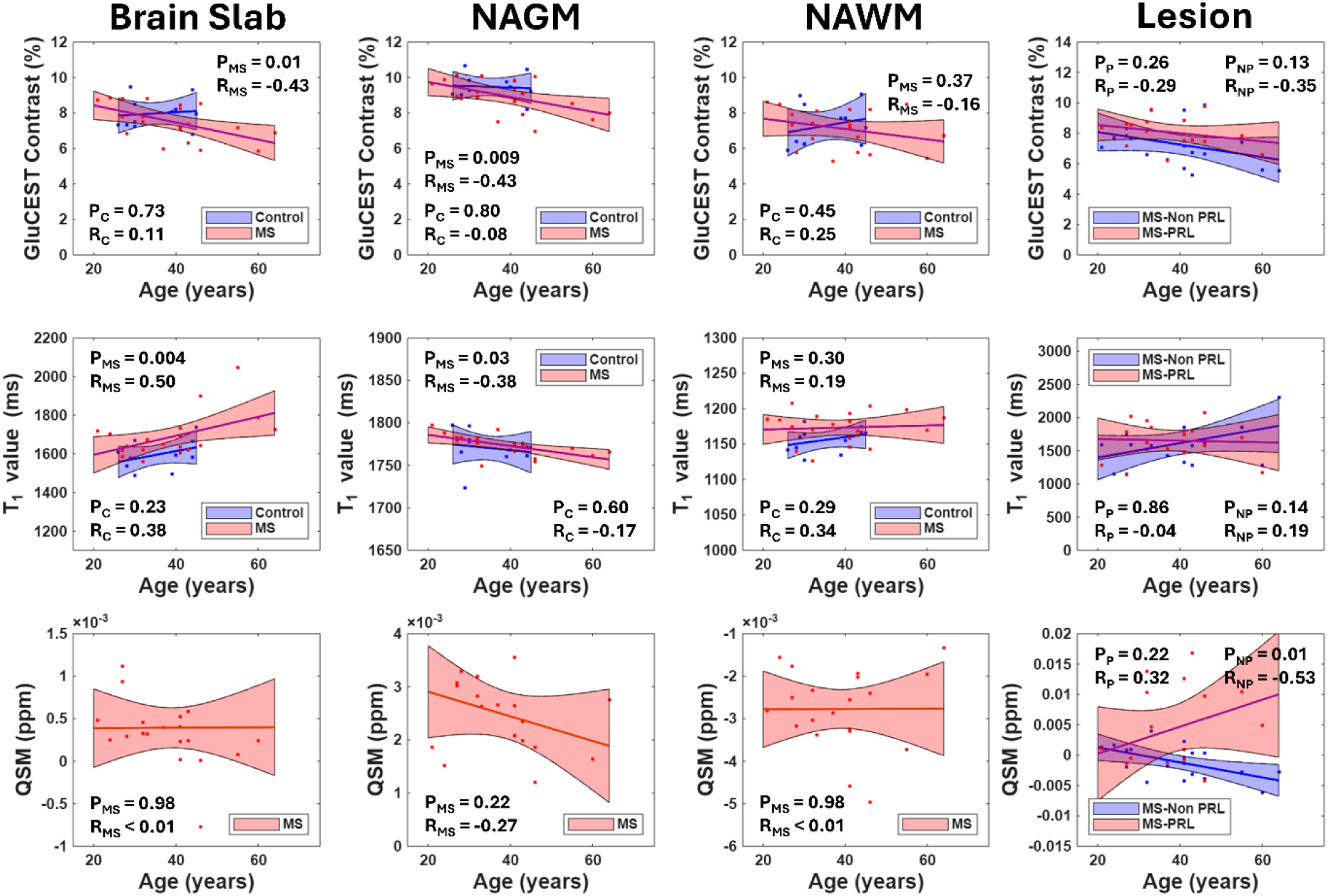
Correlations Between Age and Quantitative MRI Contrast Values. A series of linear fit plots (including shaded CI region) with displayed P-values and Pearson’s correlation coefficients for MS and healthy participant age between both GluCEST contrast and T_1_ values. Correlations with QSM values can also be seen exclusively for MS participants. (**First Row**) GluCEST contrast values can be seen plotted with MS brain slab (*P* = 0.01, t-value = -2.56) and MS NAGM (*P* = 0.009, t-value = -2.79) both having significant negative correlations. (**Second Row**) T_1_ values can be seen plotted with MS brain slab (*P* = 0.004, t-value = 3.17) having a significant positive correlation and MS NAGM (*P* = 0.03, t-value = -2.27) having a significant negative correlation. (**Third Row**) QSM values can be seen plotted with non-PRLs (*P* = 0.01, t-value = -2.58) having a significant negative correlation. Segmented PRLs and non-PRLs indicated by the subscript P and NP respectively.

Figure 5 shows GluCEST contrast, T_1_ relaxation times, and QSM value correlations with PRL and non-PRL whole brain lesion volumes in MS participants. For the GluCEST contrast values, a negative correlation is seen between non-PRL volume and NAGM (R^2^ = 0.21, *P* = 0.03) contrast. For the T_1_ relaxation times, several positive correlations are seen, including between non-PRL volume and brain slab (R^2^ = 0.42, *P* < 0.01) values as well as NAWM (R^2^ = 0.20, *P* = 0.04) values. For QSM images several negative correlations are present, including non-PRL volume and brain slab (R^2^ = 0.42, *P* < 0.01) as well as NAWM (R^2^ = 0.54, *P* < 0.01). Several negative trends are also observed including between non-PRL volume and NAGM (R^2^ = 0.18, *P* = 0.05) as well as between PRL volume and NAWM (R^2^ = 0.17, *P* = 0.06). The multiple linear regression model between lesion type (PRL vs. non-PRL) volume, and their interaction was significant (*P* = 0.048). Lesion type was a significant predictor of GluCEST signal (β = -1.24, *P* = 0.03). Conversely, neither lesion volume (β = 0.01, *P* = 0.82) nor the interaction between lesion type and volume (β = -0.0006, *P* = 0.98) significantly contributed to the model. The results from this model indicate that GluCEST signal differences between PRLs and non-PRLs are not lesion volume dependent. Correlations between brain slab and segmented GluCEST contrast and T_1_ relaxation times show no relationships except in the case of segmented HC gray matter (R^2^ = 0.36, *P* = 0.04).

**Figure 5.**
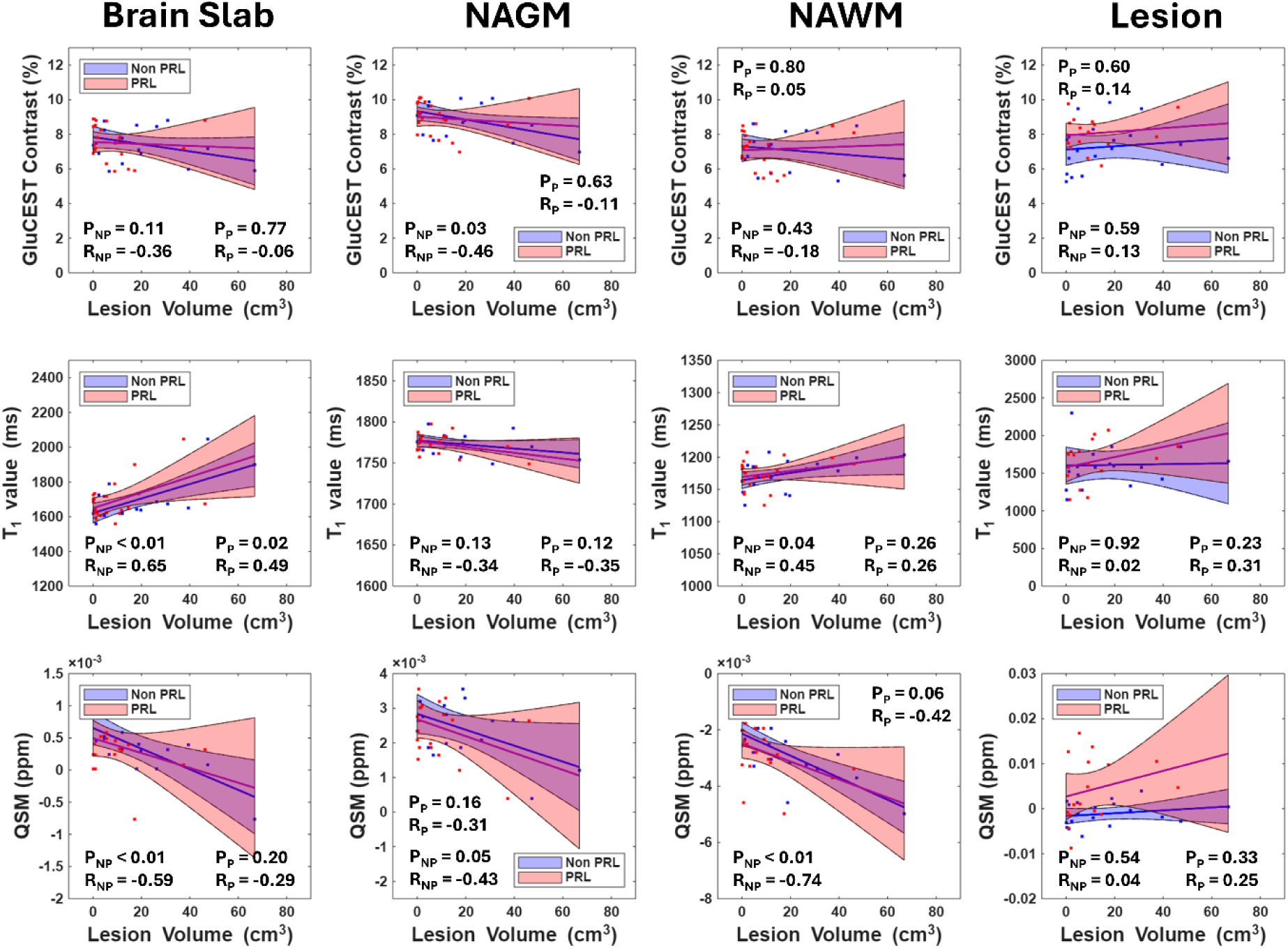
Correlations Between Lesion Volumes and Quantitative MRI Contrast Values. A series of linear fit plots (including shaded CI region) with displayed P-values and Pearson’s correlation coefficients for MS lesion subtype volume between MS GluCEST contrast, T_1_ values, and QSM values. (**First Row**) GluCEST contrast values can be seen plotted with NAGM (*P* = 0.03, t-value = -2.23) having a significant negative correlation with non-PRL volume. (**Second Row**) T_1_ values can be seen plotted with brain slab having a significant positive correlation for both PRL (*P* = 0.02, t-value = 2.39) and non-PRL (*P* = 0.001, t-value = 3.71) volume as well as NAWM (*P* = 0.04, t-value = 2.19) having a significant positive correlation with non-PRL volume. (**Third Row**) QSM values can be seen plotted with slab brain (*P* < 0.01, t-value = -3.14) and NAWM (*P* < 0.01, t-value = -4.77) having significant negative correlations to non-PRL volume. Segmented PRLs and non-PRLs indicated by the subscript P and NP respectively.

To test whether QSM contrast (and by extension iron) had any influence on resulting GluCEST contrast, correlations were made between the two segmented contrast values. The results showed that segmented GluCEST contrast and QSM values had no significant relationship. These plots can be seen as Supplemental Figure 3 and Supplemental Figure 4.

### Clinical Disability Measures are Negatively Correlated with Diffuse and PRL Tissue GluCEST

Figure 6 shows GluCEST contrast, T_1_ value, and QSM value correlations to T25FW in MS participants. For GluCEST contrast values several negative correlations are seen including for whole brain (R^2^ = 0.29, *P* = 0.03), NAGM (R^2^ = 0.27, *P* = 0.04), NAWM (R^2^ = 0.31, *P* = 0.03), and PRLs (R^2^ = 0.50, *P* < 0.01). A negative GluCEST trend is also present for non-PRLs (R^2^ = 0.24, *P* = 0.07). For T_1_ relaxation times, apart from a negative trend for PRLs (R^2^ = 0.26, *P* = 0.08), no correlations to T25FW are present. QSM values show no correlation to T25FW. Figure 7 shows GluCEST contrast, T_1_ values, and QSM value correlations to 9-HPT in MS participants. For GluCEST contrast values, again, several negative correlations are present including NAGM (R^2^ = 0.31, *P* = 0.02), NAWM (R^2^ = 0.26, *P* = 0.04), and PRLs (R^2^ = 0.54, *P* < 0.01). A negative GluCEST trend is also seen for brain slab (R^2^ = 0.25, *P* = 0.05) values as well. T_1_ values and QSM values both show no correlations to 9-HPT.

**Figure 6.**
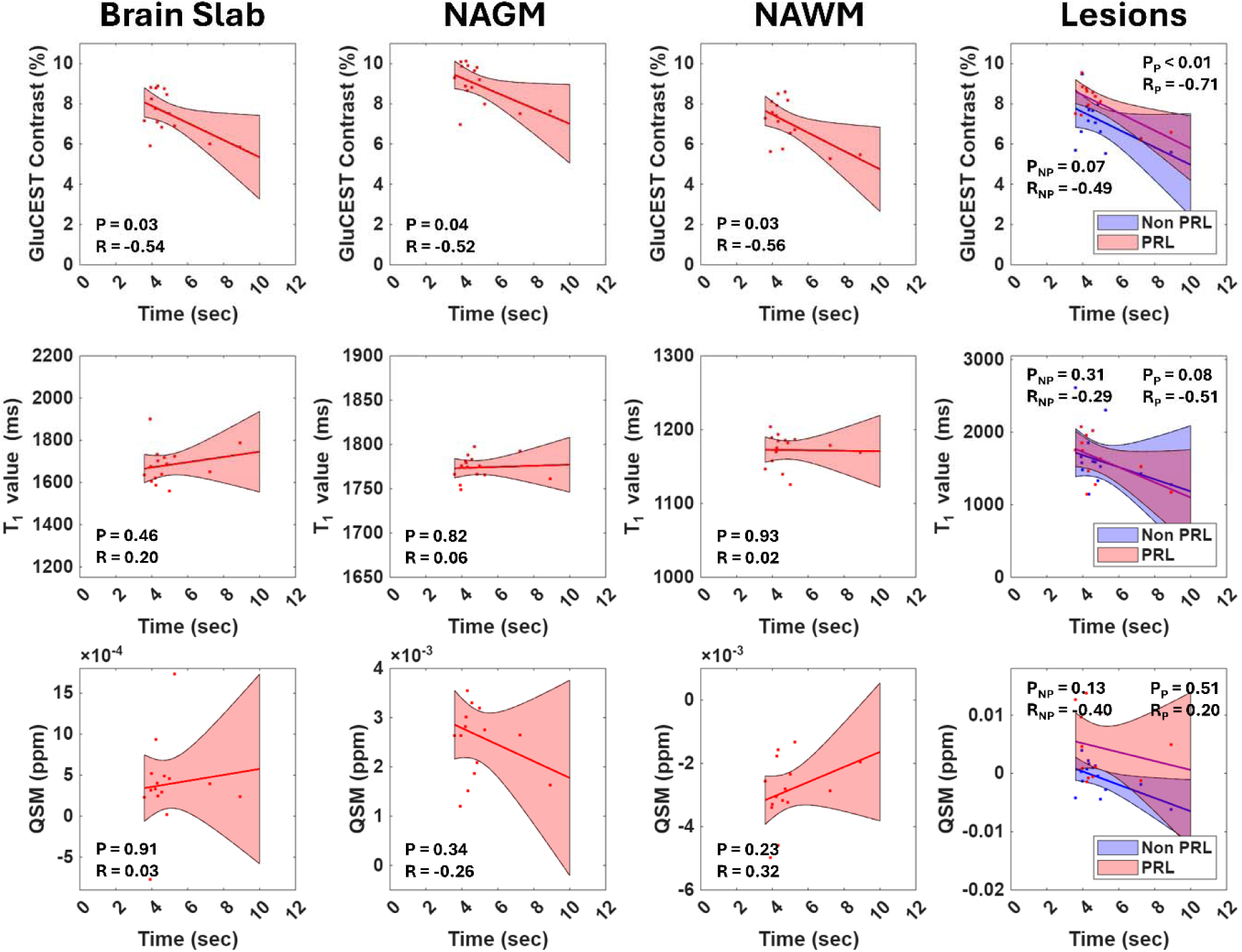
Correlations between T25FW and Quantitative MRI Contrast Values. A series of linear fit plots (incl ding shaded CI region) with displayed P-values and Pearson’s correlation coefficients between MS participant timed 25-foot walk (T25FW) for GluCEST contrast, T_1_ values, and QSM values. (**First Row**) GluCEST contrast values can be seen plotted with brain slab (*P* = 0.03, t-value = -2.34), NAGM (*P* = 0.04, t-value = -2.22), NAWM (*P* = 0.03, t-value = - 2.48), and PRLs (*P* < 0.01, t-value = -3.26) all having significant negative correlations. (**Second Row**) T_1_ values can be seen plotted with no significant correlations observed. (**Third Row**) QSM values can be seen plotted with no significant correlations observed. Segmented PRLs and non-PRLs indicated by the subscript P and NP respectively.

**Figure 7.**
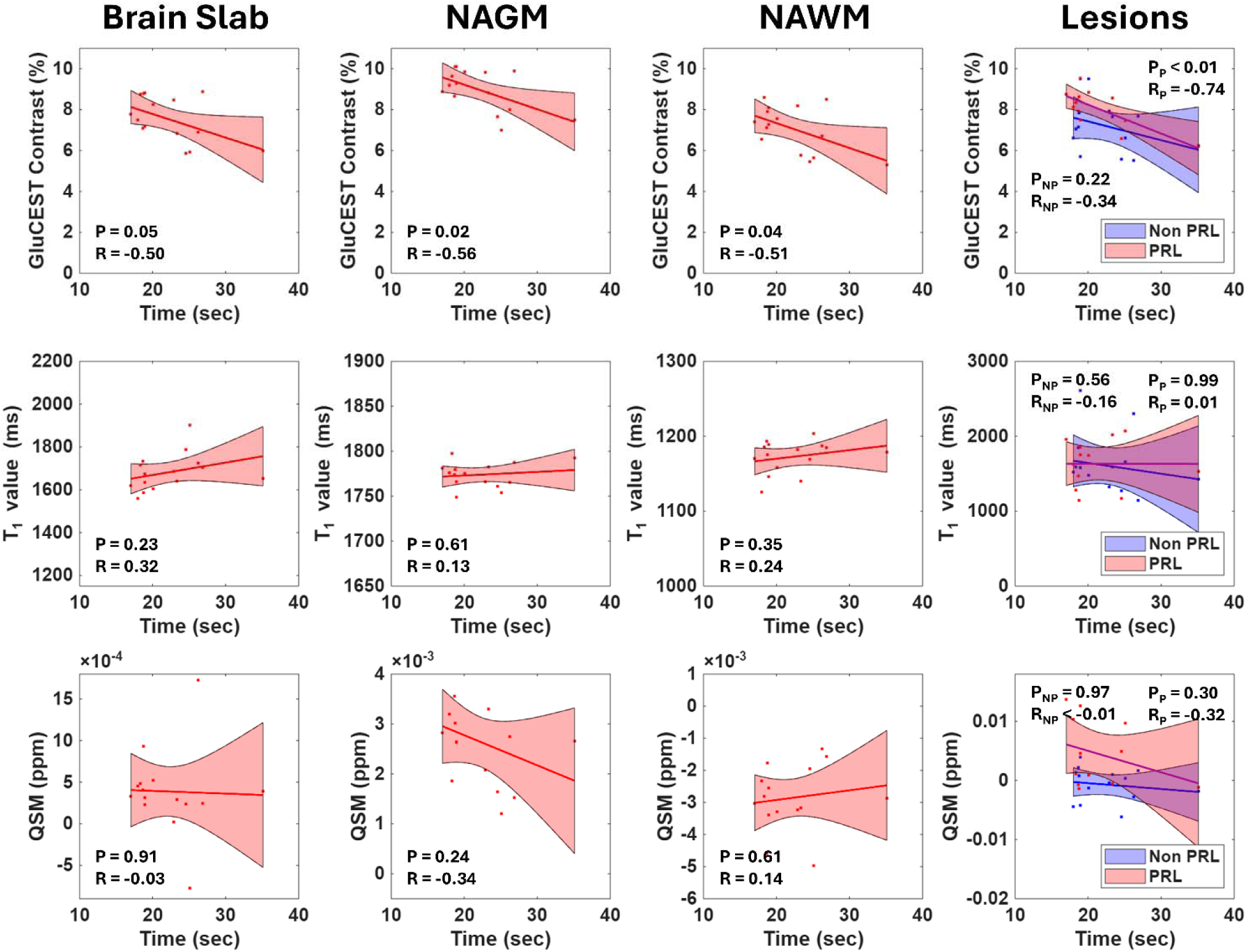
Correlations Between 9-HPT and Quantitative MRI Contrast Values. A series of linear fit plots (incl ding shaded CI region) with displayed P-values and Pearson’s correlation coefficients between MS participant 9-hole peg test (9-HPT) for GluCEST contrast, T_1_ values, and QSM values. (**First Row**) GluCEST contrast values can be seen plotted with NAGM (*P* = 0.02, t-value = -2.52), NAWM (*P* = 0.04, t-value = -2.22), and PRLs (*P* < 0.01, t-value = -3.51) all having significant negative correlations. (**Second Row**) T_1_ values can be seen plotted with no significant correlations observed. (**Third Row**) QSM values can be seen plotted with no significant correlations observed. Segmented PRLs and non-PRLs indicated by the subscript P and NP respectively.

## DISCUSSION

GluCEST contrast, averaged across PRLs, was 10.7% greater than in non-PRLs (Figure 3). It was also observed that non-PRLs were not distinguishable from NAWM. When comparing normal appearing tissue between multiple sclerosis and healthy controls, no significant differences were observed in GluCEST contrast. For PwMS, a statistically significant increase in GluCEST contrast was observed in PRLs compared to NAWM. Our findings are consistent with Srinivasan et al.^3^ who found an elevation in Glx in acute multiple sclerosis lesions using ^1^H MRS at 3T, however, in their work a difference between PRLs and non-PRLs was not investigated. Our observation of elevated levels of GluCEST contrast within PRLs compared to non-PRLs potentially supports the hypothesis that glutamatergic excitotoxicity, possibly mediated by chronic inflammation within PRLs, may be a factor linking the increased neurodegeneration and clinical disability progression reported in patients with increased PRL burden^4,37^. Prior research suggests that glutamate levels are transiently higher in acute lesions compared to healthy individuals which could be due to increased glutamate production via dysregulation of the glutamate/glutamine cycle and/or released from immune cells^38^. Excess glutamate can induce excitotoxicity which can drive progressive axonal injury. In the case of PRLs, one proposed mechanism is that iron and myelin debris within microglia/macrophages accumulate at the edge of the lesion, which could lead to increased neuronal cell death via an increase in reactive oxygen species (ROS), resulting in a further release of axonal glutamate as well as increased production from microglia and macrophages and decreased recycling from oligodendrocytes^4,10^. In combination, these processes may contribute to glutamate-spill over and increased binding to extra synaptic NMDA receptors leading to increased Ca^2+^ influx^5,39^. The increased Ca^2+^ contributes to axonal injury and neurodegeneration by prolonging plasma and mitochondrial membrane depolarization^40^.

Previously O’Grady et al.^21^ also performed GluCEST imaging at 7T and found increased contrast in the prefrontal cortex of NAGM of multiple sclerosis patients compared to healthy controls. However, in a follow up study by the same group, no difference in GluCEST contrast was observed in NAGM of multiple sclerosis patients compared to healthy controls^22^. This seeming inconsistency may be due to the different RF saturation parameters used. Furthermore, these and other previous MRS studies^3,16–18^ did not classify and compare between PRLs and non-PRLs during analyses.

A significant negative correlation between age and brain slab GluCEST contrast was observed in multiple sclerosis participants, primarily driven by changes in NAGM. A previous study by Mulhert et al.^41^ on a cohort of PwMS suggested that increased cortical atrophy resulted in decreased glutamate in NAGM, which may relate to cognitive decline. In the present work, similar correlations and trends were observed between GluCEST and disease duration in both brain slab and NAGM tissues. A negative correlation between NAGM GluCEST contrast and non-PRL volume was also observed, which also suggests potential involvement of cortical atrophy driving gray matter GluCEST changes.

Significant increases in T_1_ values were also observed in the multiple sclerosis participants compared to healthy controls in the brain slab as well as in PRLs and non-PRLs compared to NAWM and healthy control white matter. Increases in lesion T_1_ values compared to surrounding NAWM are expected due to an increase in water content, as described in previous studies^42,43^. This elevation in lesion T_1_ values, however, did not extend to a PRL and non-PRL tissue difference. This highlights that in spite of lesion T_1_ values remaining constant between PRLs and non-PRLs, the GluCEST contrast showed opposite contrasts between lesion types. This suggests that GluCEST could potentially capture metabolic information unique to the underlying pathobiology of PRLs that the T_1_ values cannot. Contrary to our results, elevated PRL T_1_ values were observed in previous work^44,45^. This difference may be due to the small sample size or the present work being composed of a single time point without knowing how old the PRLs are^46^. A positive correlation was observed between brain slab T_1_ and age as well as a negative correlation in NAGM and age for multiple sclerosis participants. Positive lesion volume correlations were observed in whole brain (for both PRL and non-PRL) and in white matter (non-PRL). Previous work^47–49^ found correlations between lesion T_1_ value and similar disease severity metrics such as disease duration and EDSS. Additionally, when investigating the relationship between T_1_ values and GluCEST contrast, correlations between the two (Supplemental Figure 1) showed no significant relationship in multiple sclerosis participants and only one correlation in the gray matter of healthy controls.

QSM values also showed a statistically significant increase in PRLs compared to non-PRLs, which matches previously reported trends in PRL tissue using QSM as a technique^50–53^. Previous work by Elkady et al.^54^ used 3T to measure, among many metrics, *in vivo* QSM values. The authors observed a significant increase over 96 weeks (21±1.3ppb) in PRLs compared to non-PRLs which are consistent with our results. This may reflect a higher concentration of iron rich myeloid cells at the lesion rim and/or demyelination in the lesion core^55^. Other MRI/histopathology studies have also confirmed the presence of iron and iron related proteins at the rim of chronic active lesions both *in vivo* and post-mortem^56,57^. From our results, we also observed no correlation between the GluCEST and QSM measurements in any of the tissue classes examined. This lack of correlation suggests that endogenous iron may not be a major driving factor of the GluCEST signal and supports our claim that glutamate is the primary driver of the observed relative changes. QSM and GluCEST images may also provide complementary information about different aspects of the disease which are limited by when they are acquired. However, these claims need to be further evaluated via a longitudinal study into PRL evolution.

Regarding clinical metric correlations, our results show that multiple sclerosis participants who had lower GluCEST contrast values diffusely, performed worse in both the T25FW and 9-HPT. Additionally, these correlations were observed in GluCEST contrast in PRLs but were absent in non-PRLs, suggesting that PRL tissue may be uniquely involved in this disease process. Our work also aligns with prior studies, one example being from Klineova et al.^58^ who observed a negative association between gray and white matter volumes and slower T25FW times. Other studies, such as those by Kazimuddin et al.^59^ and Cordani et al.^60^, have also shown that worse performance on 9-HPT was associated with multiple sclerosis patients who had PRLs with lower fractional anisotropy values. However, these findings should be interpreted cautiously as further experiments are needed to investigate associations between these imaging biomarkers and other clinical measures of disability, such as the symbol digit modalities test (SDMT) and EDSS.

Limitations of our work include having a relatively small cohort size, which could lead to potential false positives as well as not being able to perform disease subgroup analyses. Additionally, as mentioned previously, having only a single time point for each participant precludes investigations into time-dependent changes in GluCEST contrast. Being a single center study and the lack of gadolinium contrast to distinguish between contrast enhancing and non-contrast enhancing lesions at the time of 7T MRI is also another limiting factor of the present work. Additionally, the GluCEST acquisition is currently only able to reliably cover a volumetric slab region of 20 mm and therefore investigations into whole brain contrast changes cannot be made. Additionally, the GluCEST signal may be influenced by other factors such as contributions from other macromolecular groups and as such the results should be interpreted cautiously. When it comes to the PRL and non-PRL segmentations, inter-rater reliability or reproducibility metrics were also not considered in this work and should be included in future studies. Future studies incorporating additional time points on the same participants could provide predictive information about the progression of tissue injury. Lastly, additional downfield MRS measurements^61–63^ would allow for correlating current CEST-based metrics with molecules (i.e., ATP, NAD+, Tryptophan) that cannot be directly investigated via CEST.

In conclusion, GluCEST imaging at 7Tshows a 10.7% elevation in GluCEST contrast within PRLs compared to non-PRLs. Further analysis showed that this contrast difference was independent of any lesion volume effects. However, it should be noted that we are unable to comment on the effects of volume at the level of individual lesions. Significant correlations between GluCEST contrast and clinical disability metrics (T25FW & 9-HPT), potentially suggest glutamate dysregulation, which may be a factor linking PRL-related chronic inflammation with neurodegeneration. GluCEST imaging provides unique metabolic information in characterizing PRLs that is not captured by conventional structural imaging, T_1_ mapping, or susceptibility imaging. Thus, GluCEST may help in probing the underlying mechanisms associated with more severe clinical outcomes. These findings suggest that metabolic imaging may serve as an important tool for stratifying patients at risk for greater neurodegeneration and testing therapies targeting excitotoxic pathways. The preliminary nature of these results allows for the generation of novel hypotheses which primes future work incorporating longitudinal tracking and advanced MR sequences specific to other important molecular species that may help to identify potential targets to intervene and to combat neurodegeneration in multiple sclerosis.

## DATA AVAILABILITY

Data is available to investigators upon reasonable request.

## AUTHOR CONTRIBUTIONS

P.S.J., M.K.S., R.R. contributed to conception and design of the study; P.S.J., B.S., N.B., N.E.W., D.R., A.E., M.K.S., R.R., R.T.S, M.M.A., A.S. contributed to the acquisition and analysis of data; P.S.J., B.S., N.B., N.E.W., A.E., R.T.S, M.M.A., A.S., L.Y., S.T., A.B.O., J.D., D.R., M.K.S., R.R. contributed to drafting the text of preparing the figures

