## Supplemental Figures and Tables for "Altered Glutamate Homeostasis in Paramagnetic Rim Lesions of Patients with Multiple Sclerosis"

**Supplemental Material**

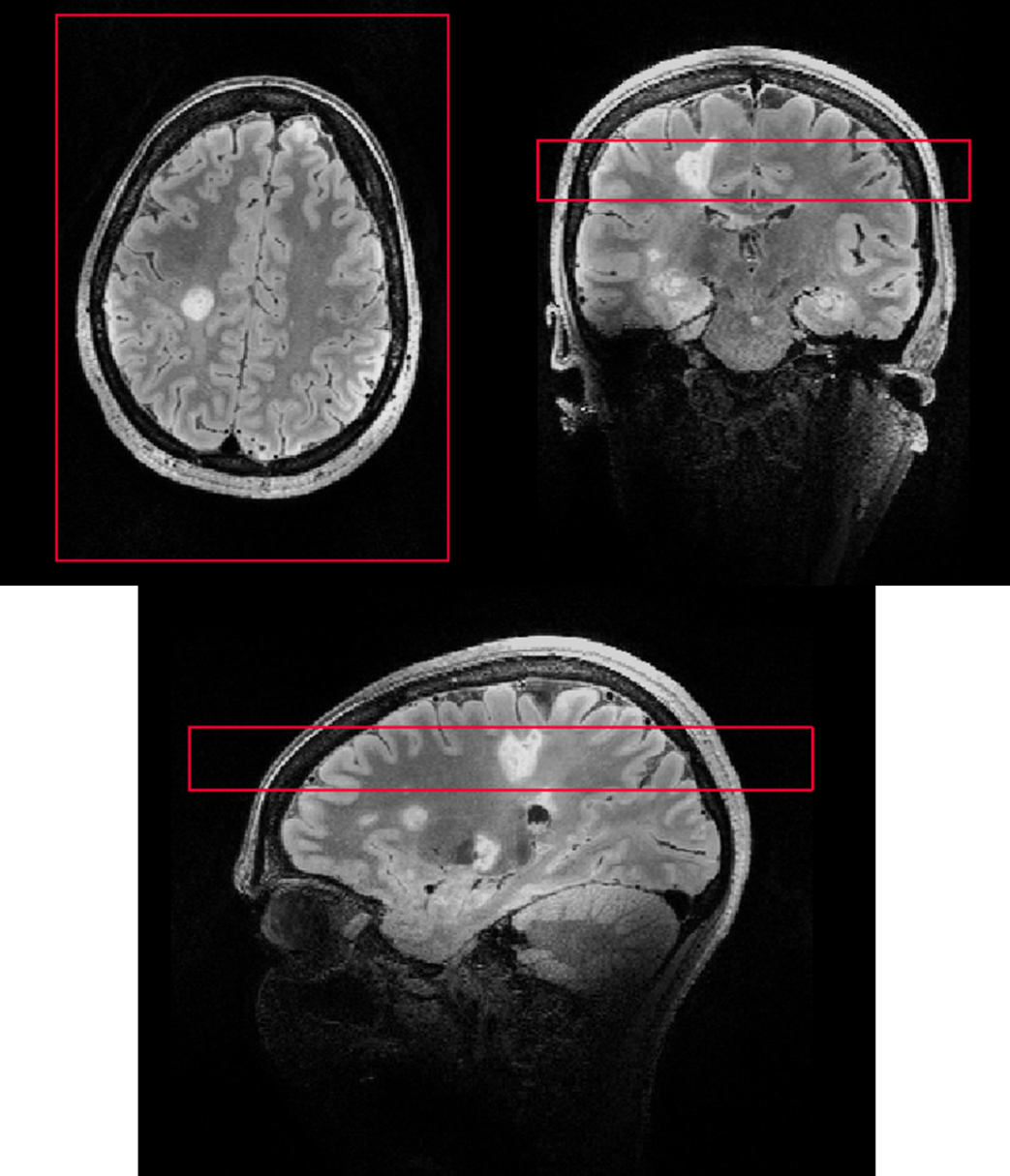

**Supplemental Figure 1.** A set of representative example images of the brain slab volume in which axial (top left), coronal (top right), and sagittal (bottom center) FLAIR images can be seen with an overlaid red outline corresponding to the size and location of the 20 mm GluCEST “brain slab” acquisition.

|  | | **T_1_ value (ms)** | **QSM (ppb)** | **GluCEST (%)** |
| --- | --- | --- | --- | --- |
| **Whole Brain** | **Control** | 1593.9±75.4 | - | 7.97±0.85 |
|  | **MS** | 1688.6±114.5 | 0.39±0.48 | 7.50±1.01 |
| **Gray Matter** | **Control** | 1770.3±19.9 | - | 9.47±0.68 |
|  | **MS** | 1773±12.5 | 2.46±0.95 | 8.94±0.94 |
| **White Matter** | **Control** | 1155.9±17.1 | - | 7.27±1.18 |
|  | **MS** | 1173.4±22.2 | -2.77±0.94 | 7.12±1.09 |
| **Non PRL** | | 1609.2±365.6 | -0.68±3.25 | 7.27±1.36 |
| **PRL** | | 1653.0±280.9 | 4.26±7.27 | 8.05±0.97 |

**Supplemental Table 1.** **Summarized groupwise T_1_ values, QSM values, and GluCEST contrast values**

Summarized average (standard deviation) values for T_1_ and GluCEST contrast values across all segmented tissue types for both MS and healthy control data.

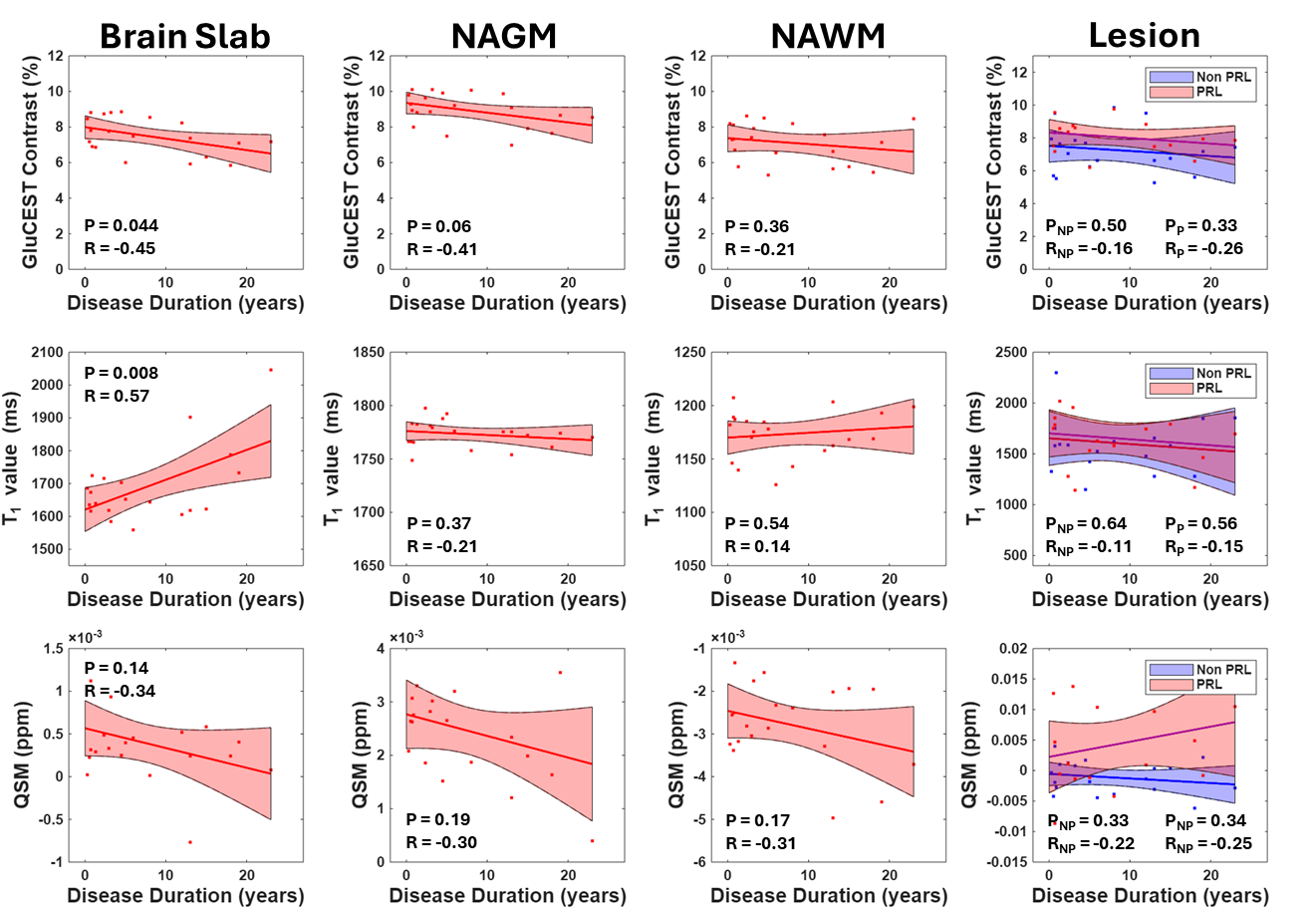

**Supplemental Figure 2.** A series of linear fit plots (including shaded CI region) with displayed P-values and Pearson’s correlation coefficients for disease duration between MS GluCEST contrast, T_1_ values, and QSM values. (**First Row**) GluCEST contrast values can be seen plotted with brain slab (*P* = 0.04, t-value = -2.16) having a significant negative correlation. (**Second Row**) T_1_ values can be seen plotted with brain slab (*P* = 0.008, t-value = 2.93) having a significant positive correlation. (**Third Row**) QSM values can be seen plotted with no significant correlations observed. Segmented PRLs and non-PRLs indicated by the subscript P and NP respectively.

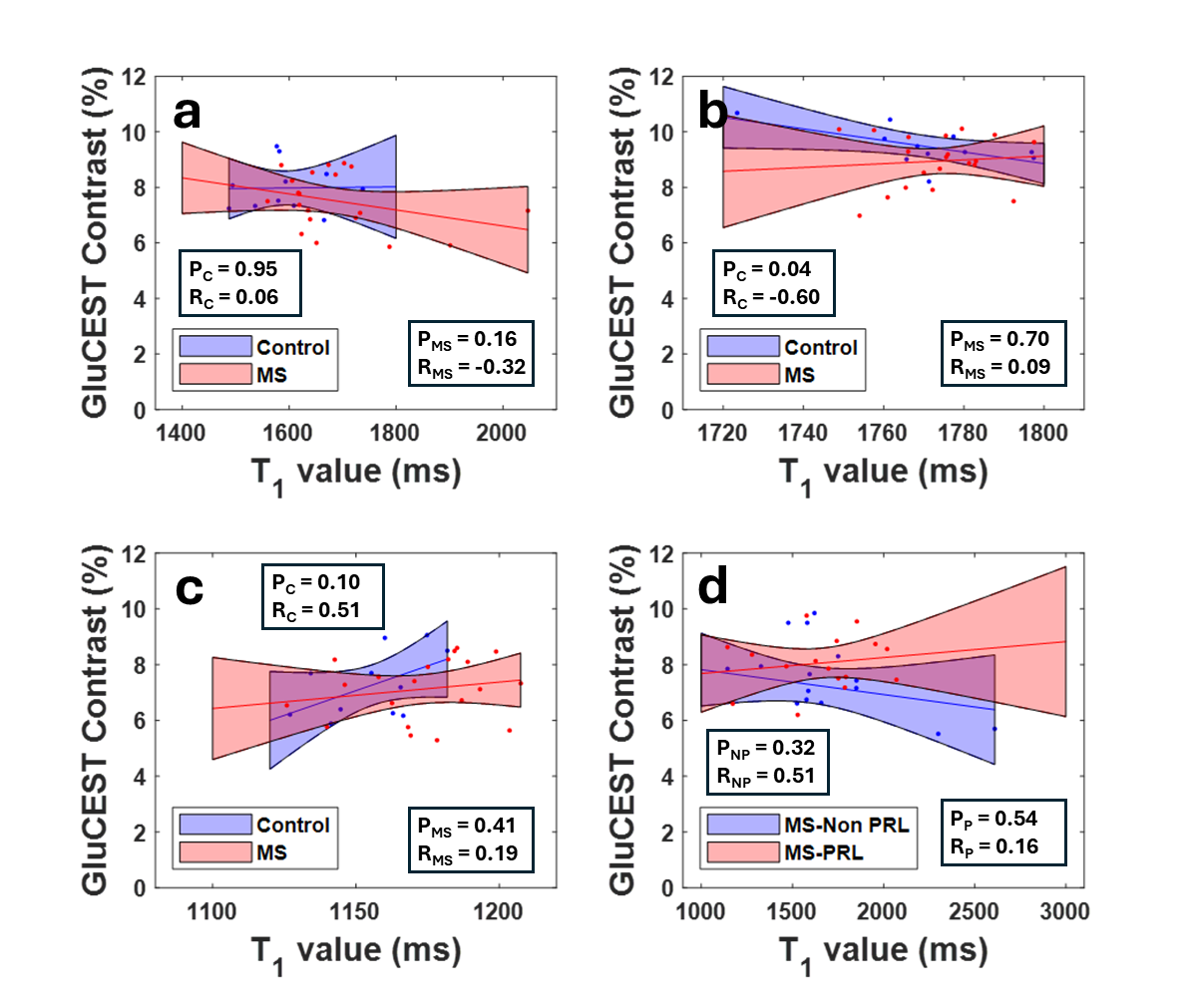

**Supplemental Figure 3.** Correlations between T_1_ and GluCEST contrast values for each of the tissue regions including (a) whole brain slab, (b) gray matter, (c) white matter, and (d) PRL and Non PRL tissue. The only correlation that was observed was in healthy control diffuse gray matter tissue (*P* = 0.04, t-value = -2.28, R = -0.60).

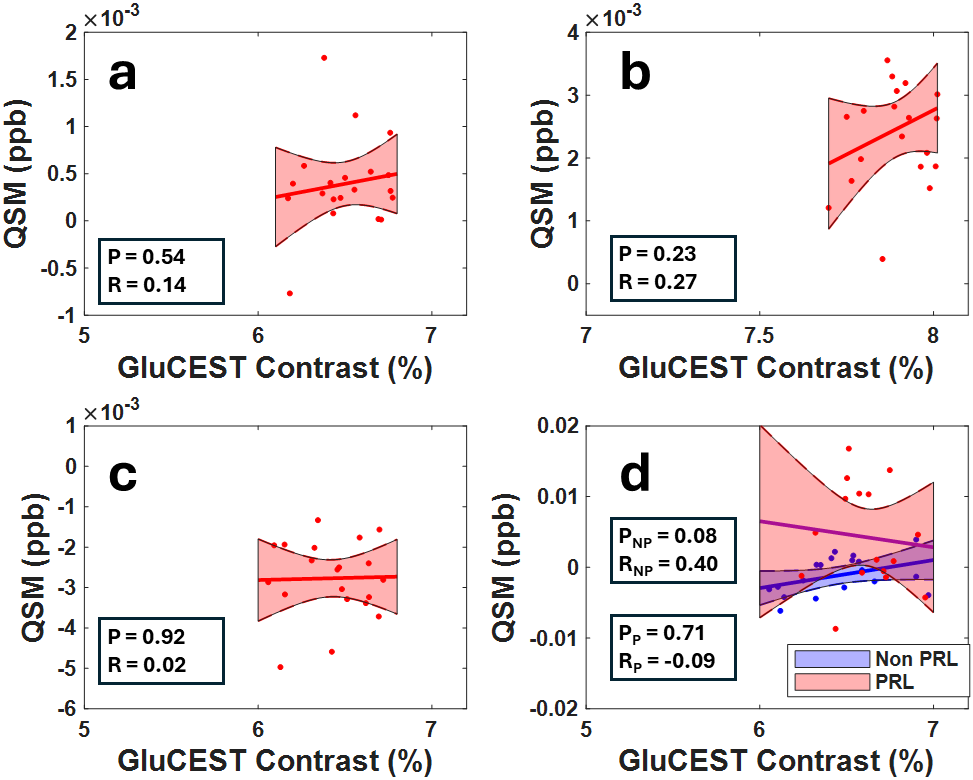

**Supplemental Figure 4.** Correlations between GluCEST contrast and QSM values for each of the tissue regions including (a) whole brain slab, (b) gray matter, (c) white matter, and (d) PRL and Non-PRL tissue. No correlations were observed in any of the tissue regions.
